# Vestibular Perception, Balance Impairment, and Fall Risk in Community-Dwelling Older Adults

**DOI:** 10.64898/2026.02.19.26346653

**Authors:** Yuxiao Li, Zaeem Hadi, Rebecca M Smith, Barry M Seemungal, Toby J Ellmers

**Affiliations:** Centre for Vestibular Neurology, Department of Brain Sciences, Imperial College London, UK

**Keywords:** vestibular agnosia, vestibular perceptual thresholds, vestibulo-ocular reflex, postural sway, fall risk

## Abstract

**Background:** Vestibular complaints are common in older adults and are linked to imbalance and falls. Some older adults show impaired vestibular perception despite preserved peripheral-reflex (“vestibular agnosia”). Yet it remains unclear if vestibular agnosia is independently linked to imbalance and falls in otherwise healthy older adults. We therefore investigated the prevalence of vestibular agnosia in community-dwelling older adults, and examined its association to balance and prospective falls.

**Methods:** Vestibular perceptual thresholds were measured during yaw-plane rotational chair testing. Postural sway and instrumented Timed-Up-and-Go were assessed using wearable sensors, and falls were recorded prospectively over six-month. Vestibular agnosia was identified using K-means clustering. Multivariable regressions examined associations between perceptual thresholds and balance outcomes; logistic and negative binomial regressions evaluated associations with prospective falls.

**Results:** Among 166 participants (75.4 years; 81.9% female), 18.7% were classified as having vestibular agnosia. These individuals had worse cognition and somatosensation. Elevated (i.e. worse) vestibular perceptual thresholds were independently associated with greater sway velocity when standing on foam with eyes-open (adjusted β=0.002, *p*=0.03). Associations with other balance outcomes were attenuated after adjustment. Vestibular perceptual thresholds were not associated with prospective falls (odds of ≥1 fall: adjusted OR=0.99, *p*=0.65; fall counts: adjusted IRR=1.02, *p*=0.35).

**Conclusions:** Approximately one-fifth of healthy older adults exhibit vestibular agnosia. While elevated perceptual thresholds are independently associated with poorer balance, they did not predict falls. Vestibular perceptual testing provides complementary insight into age-related balance impairment, although its utility in fall-risk prediction requires further investigation.

**Key Points:**

- Approximately one-fifth of healthy older adults had vestibular agnosia (impaired vestibular perception despite intact peripheral function)
- Older adults with vestibular agnosia have poorer cognition, reduced lower limb somatosensation, and higher anxiety.
- Higher (i.e. worse) vestibular perceptual thresholds were independently associated with greater sway velocity when standing on foam (eyes open).
- Higher vestibular perceptual thresholds were only associated with slower TUG performance and greater eyes-closed foam sway in unadjusted models.
- Vestibular perceptual thresholds did not predict prospective falls over 6 months.

## INTRODUCTION

Approximately 30% of community-dwelling older adults fall annually [1,2], and age-related balance decline plays a central role [3]. Postural control integrates visual, proprioceptive, and vestibular inputs [4], with vestibular function supporting self-motion perception and spatial orientation [5]. However, vestibular perceptual thresholds, which quantify the smallest self-motion stimulus that can be reliably perceived on a given axis [6,7], tend to increase with age [8] and relate to poorer balance performance [9–13]. Recent work has also linked impaired vestibular perception to increased prospective falls in older adults with Type-2 diabetes [11]. However, evidence for age-related differences in perceptual thresholds is mixed [14], and later-life comorbidities may contribute. These mixed findings highlight the importance of motivate examining vestibular function, and its relationship with balance performance and falls, specifically in generally healthy community-dwelling older adults.

Impaired vestibular sensation can occur due to either disturbed peripheral [15] and/or central vestibular function [16,17]. The latter, whereby impaired vestibular sensation occurs despite vestibular ocular activation (indicating preserved inner ear and reflex function), is known as “vestibular agnosia” [17]. While vestibular agnosia is well-characterised in neurological conditions (e.g., traumatic brain injury [16–18] and Parkinson’s Disease [19]), age-related vestibular agnosia in otherwise healthy older adults has been less examined. However, recent evidence suggests that ∼25% of older adults who test positive for BPPV report no dizziness [20], indicating a high prevalence of vestibular agnosia in community-dwelling older adults. This has implications for falls guidelines in older adults, supporting our prior argument for incorporating objectively vestibular sensation assessment into screening beyond self-reported ‘dizziness’ [21,22].

Existing studies exploring vestibular perception in older adults did not assess peripheral vestibular function, making it difficult to determine whether perceptual deficits arise from central processing or peripheral vestibular dysfunction. Furthermore, the prevalence and risk factors of vestibular agnosia in community-dwelling older adults remain unknown. Existing research has largely focused on associations between vestibular perception and postural sway [9,10], while overlooking more ecologically valid tasks like walking and turning. This is important because most falls in older adults occur during dynamic activities, whereas quiet standing only accounts for ∼10% [23]. Turning is more vestibular-dependent than straight-line walking [24], and slower turning has been liked to increased fall risk [25].

To address these gaps, we aimed to investigate the prevalence and associated characteristics of vestibular agnosia in community-dwelling older adults; and to examine whether impaired vestibular perception relates to balance performance (both static postural sway and instrumented timed up-and-go) and 6-month prospective falls, after controlling for key covariates including peripheral dysfunction. We hypothesize that vestibular agnosia reflects age-related central vestibular processing impairment that disrupts the conscious perception of self-motion and utilization of vestibular signals in the brain. We therefore predict that vestibular agnosia will result in poorer balance (particularly in vestibular-dependent balance tasks, e.g., eyes-closed on foam and turning), and be independently associated with increased prospective fall risk.

## METHODS

### Participants and recruitment

We recruited community-dwelling older adults aged ≥60 years in London (April – December 2024). Exclusion criteria were: a clinical diagnosis of a progressive neurological disorder (e.g., Alzheimer’s, Parkinson’s, multiple sclerosis); inability to walk 10 meters unaided; severe visual or auditory impairment (i.e., legally blind/deaf); and major cognitive impairment (Montreal Cognitive Assessment (MoCA) score of ≤18 [26]), to ensure participants could understand and complete the threshold tests. In addition, 20 healthy younger adults (aged 22–32 years) without neurological/balance disorders were recruited to enable comparison with normative values for vestibulo-ocular reflex (VOR) and perceptual thresholds (see below).

All participants provided written informed consent, and the study was approved by East of England – Essex Research Ethics Committee (IRAS ID: 324021).

### Measures

### Vestibular perceptual and vestibulo-ocular reflex (VOR) thresholds

Figure 1. illustrates the apparatus used to objectively access vestibular perceptual and VOR thresholds during passive yaw-plane rotations using a vibration-free motorized rotating chair (Contraves, USA) [17,27]. Participants were seated upright with head support and safety harness, with visual and auditory cues minimised (light off, blackout curtain, and white noise), and were asked to press the button (left or right) as soon as they perceived a rotation in the respective direction.

**Figure 1.**
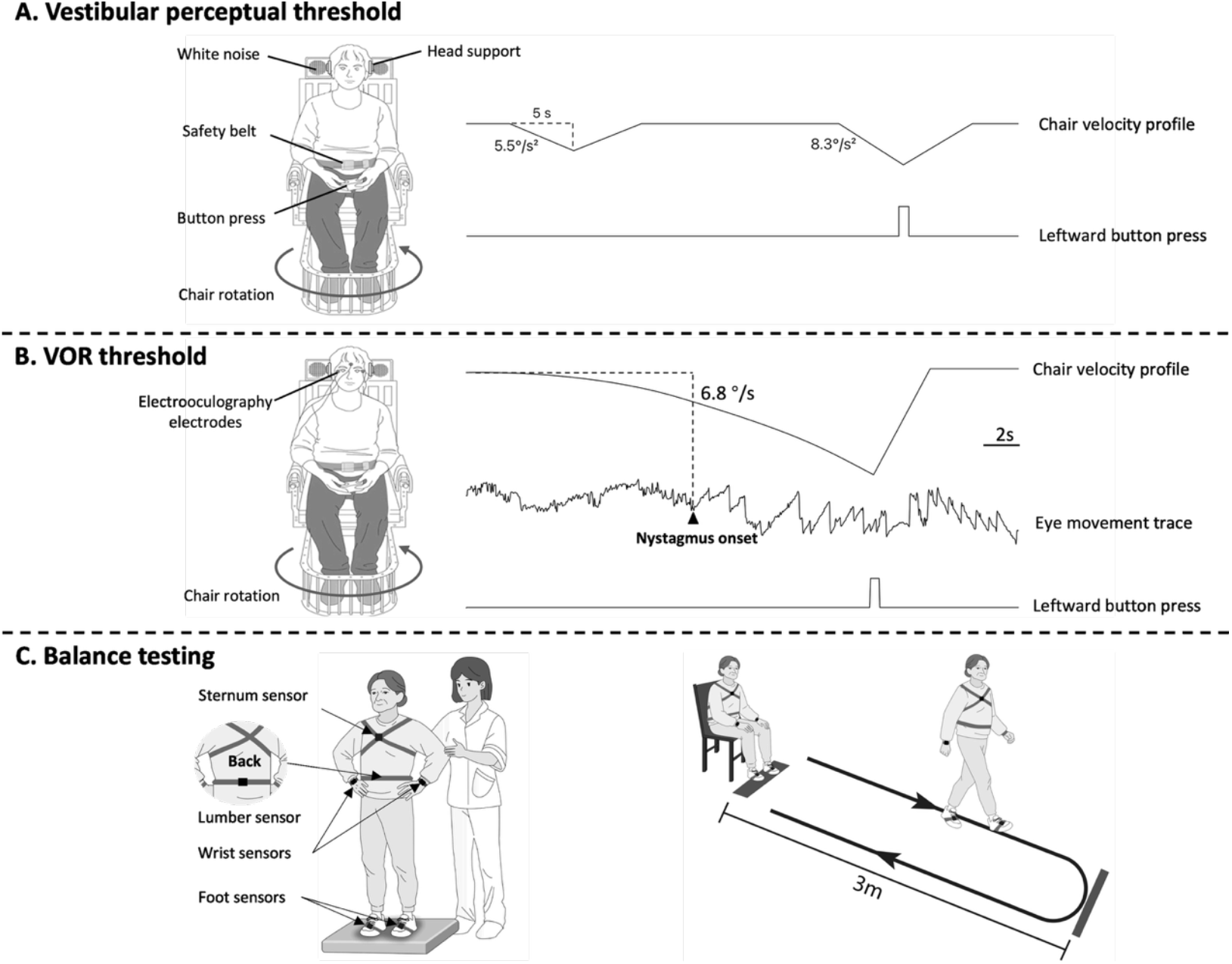
Yaw-axis rotational chair set-up and stimulus profiles for vestibular perceptual and VOR threshold measurements. **A:** Vestibular perceptual threshold paradigm. The upper trace shows the chair velocity profile during a constant-acceleration rotation (up to 5 s), with acceleration level adjusted across trials by the modified binary search (MOBS) staircase algorithm (illustrated here with 5.5°/s^2^ and 8.3°/s^2^). The lower trace indicates the timing of a leftward button press; correct directional responses within the 5-s acceleration phase were scored as correct, whereas incorrect or late responses were scored as misses. **B:** Vestibulo-ocular reflex (VOR) threshold paradigm. Yaw-plane rotations with linearly increasing acceleration (0.3°/s^2^ every 3 seconds). The upper trace shows the chair velocity profile, the middle trace the horizontal eye-movement response recorded with electrooculography, with the arrow indicating the onset of nystagmus, the lower trace indicates the timing of the participant’s button press to report perceived direction of rotation. **C:** Balance testing. Participants completed instrumented balance assessments using six inertial wearable sensors (wrists, sternum, lower lumbar, and feet). For postural sway on foam: participants stood quietly on a soft rectangular foam (50 × 41 × 6 cm, Airex®), with standardized hip-width stance and with their arms placed on their hips. For Timed Up and Go (TUG), participants were asked to stand up from a chair, walk 3 meters, turn around, return, and then seat back down.

Perceptual testing (**Figure 1A**), stimuli followed a constant acceleration profile, with acceleration determined by the modified binary search (MOBS) staircase algorithm [28] with independent randomly left-/right-ward rotations (≤5 s; 0.2 Hz; matched deceleration). VOR thresholds were obtained simultaneously during perceptual testing; but as perceptual thresholds were much higher than VOR thresholds, the algorithm rapidly moved away from the peri-VOR thresholds range. Therefore, we additionally assessed VOR thresholds separately using the same yaw-plane chair rotations with linearly increasing acceleration (0.3°/s^2^ every 3 s) [17] (**Figure 1B**). More detailed information regarding the chair and algorithm can be found in **Supplementary Methods**.

To control for potential confounding by reaction speed and vigilance on perceptual thresholds, participants completed a computerised hand reaction time task [17].

### Balance assessments

Balance was assessed using six inertial wearable sensors (Opal™, APDM, Inc., Portland, OR, USA) (**Figure 1C**). For the s*tatic balance*, analyses focused on soft-surface conditions (eyes-open and eyes-closed for 30 seconds each [29]), as previous studies in traumatic brain injury distinguished vestibular agnosia groups on foam surface but not on solid surface [17]. Sway velocity and sway area were calculated. Also, participants performed an *instrumented timed up-and-go (TUG) test* as a measure of functional balance and mobility [30]. We derived total duration, turn angle, and turn velocity, focusing TUG analyses on yaw-plane angular velocity components (turn velocity). All parameters were derived using validated algorithms within the Mobility Lab software platform [31,32].

### Prospective Falls

Participants were provided with a falls diary in which they were instructed to list any falls experienced over 6-month follow-up, with telephone and/or email at 3 and 6 months to verify entries and ascertain falls. Falls were defined according to the World Health Organization (WHO) as “an event which results in a person coming to rest inadvertently on the ground, floor or other lower level” [33].

### Demographics and health-related variables

Age, sex, ethnicity, education, hypertension and diabetes, number of medications, smoking, alcohol use, previous falls and dizziness history in last 12-month were collected by questionnaire. Validated questionnaires included Short Falls Efficacy Scale–International (FES-I) [34]; Hospital Anxiety and Depression Scale (HADS) [35], Montreal Cognitive Assessment (MoCA) [26], Trail Making Test Part B (TMT-B) [36], Dizziness Handicap Inventory (DHI) [37], and Visual Vertigo Analogue Scale (VVAS) [38]. Anthropometric data included body mass index (BMI), lower limb strength (5-times sit-to-stand test) [39], ankle vibration thresholds [40,41]. More details are provided in the **Supplementary Methods**.

### Statistical Analysis

Between-group comparisons were performed using independent *t*-tests or Mann– Whitney U tests (continuous) and χ^2^ tests (categorical). Baseline *P*-values are unadjusted and should be interpreted as exploratory. All statistical tests were two-sided, with a significance level set at p *<* 0.05. Data analyses were conducted using SPSS Statistics (version 29.0) and R software (Version 4.4.2).

We used complete-case analyses, excluding participants with missing outcomes from the Aim-specific analyses (**Figure 2**). For ***Aim 1***, we used a two-step exclusion based on younger-adult normative perceptual threshold cut-off (mean + 2 SDs: 6.94°/s), excluding older adults with both elevated perceptual (> 6.94°/s) and VOR thresholds (>9.65°/s) – defined as peripheral vestibular hypofunction [42]. Vestibular agnosia was then identified via K-means clustering [43] of left/right perceptual thresholds or age (full sample and older-only), while perceptual thresholds were modelled continuously for Aims 2–3 due to imbalanced cluster sizes. For ***Aim 2***, we used multivariable linear regression to test associations between perceptual thresholds and balance outcomes, reporting unadjusted and fully adjusted models (age, sex, MoCA, VOR thresholds, ankle vibration thresholds, and 5-times sit-to-stand). ***Aim 3*** analysed whether vestibular perceptual thresholds predicted 6-month prospective falls using logistic regression (≥1 fall; yes/no) and negative binomial regression (fall counts). We fitted unadjusted and covariate-adjusted models (VOR thresholds, age, sex, prior falls, ankle vibration thresholds, and 5-times Sit-to-Stand duration).

**Figure 2.**
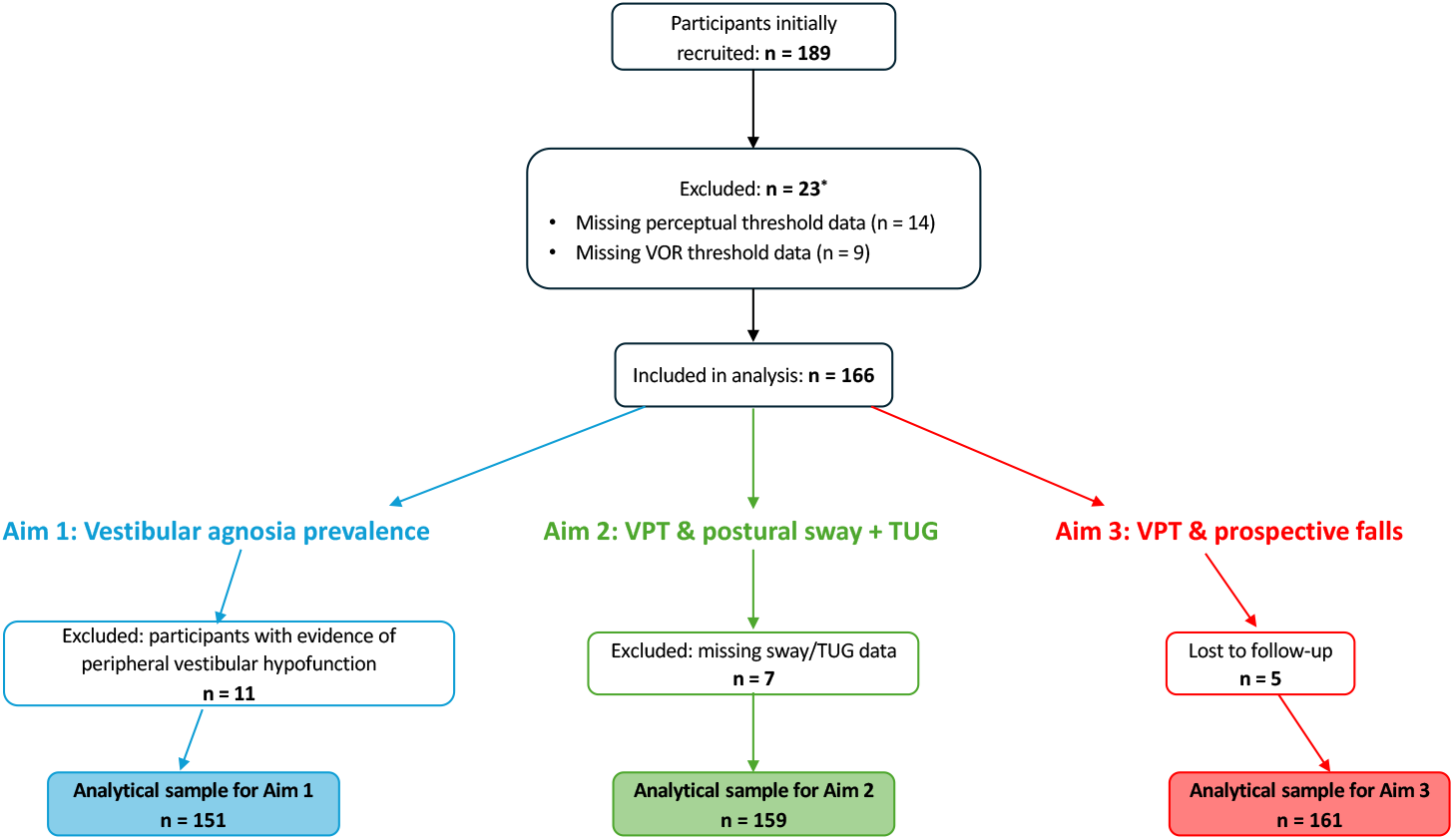
Flow of participants through research aims.

Detailed descriptions of analyses for each aim are provided in the **Supplementary Methods**.

## RESULTS

### Participant Characteristics

Of the 189 participants, 14 were excluded for missing vestibular perceptual threshold data (our primary variable) and additionally 9 for missing VOR threshold (see flow-diagram, **Figure 2**), leaving 166 participants included (**Table 1**). The mean age was 75.4±6.2 years (range 62–90), 81.9% were female, and 43.3% reported falling in the past year. The median vestibular perceptual and VOR thresholds were 8.6 °/s and 4.14 °/s, respectively.

**Table 1.** Baseline Characteristics of Participants.

| Variable | All*<br>(n=166) | VA- group<br>(n = 126; 81.3%) | VA+ group<br>(n = 29; 18.7%) | p |
| --- | --- | --- | --- | --- |
| **Demographics** |  |  |  |  |
| Age (years), mean ± SD | 75.4±6.2 | 75.0±6.3 | 76.5±6.2 | 0.23 |
| Female sex, n (%) | 136 (81.9%) | 104 (82.5%) | 25 (86.2%) | 0.84 |
| Body mass index (kg/m<sup>2</sup>), mean ± SD | 25.55±4.15 | 25.72±4.05 | 25.38±4.79 | 0.69 |
| Ethnicity: White, n (%) | 152 (91.6%) | 116 (92.1%) | 25 (86.2%) | 0.53 |
| Educational level, n (%) |  |  |  | 0.37 |
| Non-tertiary | 77 (46.4%) | 58 (46.0%) | 16 (55.2%) | - |
| University or higher | 89 (53.6%) | 68 (54.0%) | 13 (44.8%) | - |
| **Sensory and Vestibular Function** |  |  |  |  |
| Perceptual threshold (°/ s), median (IQR) | 8.60 (4.95, 12.73) | 6.28 (4.45, 9.70) | 20.40 (16.13, 24.45) | **<0.001** |
| VOR threshold (°/ s), median (IQR) | 4.14 (2.46, 6.46) | 3.31 (2.24, 5.55) | 5.40 (3.76, 7.08) | **0.006** |
| Hand reaction time (s), median (IQR) | 0.51 (0.45, 0.59) | 0.52 (0.45, 0.57) | 0.52 (0.45, 0.63) | 0.89 |
| Ankle vibration threshold, median (IQR) | 6.5 (4.7, 8.0) | 6.6 (5, 8) | 6.0 (3.8, 7.5) | **0.02** |
| **Medical History** |  |  |  |  |
| Hypertension, n (%) | 51 (30.7%) | 40 (31.7%) | 10 (34.5%) | 0.78 |
| Diabetes, n (%) | 5 (3.0%) | 4 (3.2%) | 1 (3.4%) | 0.94 |
| No. medications ≥4, n (%) | 37 (22.3%) | 26 (20.6%) | 9 (31.0%) | 0.23 |
| Smoking history (smoke or quit), n (%) | 80 (48.2%) | 61 (48.4%) | 15 (51.7%) | 0.75 |
| Alcohol use, n (%) | 117 (70.5%) | 92 (73.0%) | 18 (62.1%) | 0.24 |
| **Fall history** |  |  |  |  |
| Past 12-month fall history, n (%) | 71 (43.3%) | 50 (40.0%) | 13 (46.4%) | 0.53 |
| Short FES-I, median (IQR) | 9 (8, 11) | 9 (7, 11) | 10 (8, 12) | 0.09 |
| **Physical and psychological Function** |  |  |  |  |
| 5 Sit-to-Stand duration (s), median (IQR) | 16.70 (13.86, 20.07) | 16.03 (13.61, 19.25) | 17.89 (15.89, 21.49) | **0.03** |
| TUG duration (s), median (IQR) | 12.69 (10.60, 15.04) | 12.31 (10.36, 14.54) | 12.98 (11.56, 15.25) | 0.21 |
| MoCA, median (IQR) | 28 (27, 29) | 28 (27, 29) | 27 (24.5, 28.5) | **0.01** |
| TMT-B, median (IQR) | 94 (74, 120) | 90 (70, 117) | 100 (82, 132) | 0.16 |
| HADS-A, median (IQR) | 3 (1, 7) | 3 (1, 5) | 5 (2, 8) | **0.04** |
| HADS-D, median (IQR) | 2 (1, 4) | 2 (1, 4) | 3 (1, 6) | 0.38 |

\*All includes 166 participants. However, 11 participants met criteria for peripheral vestibular hypofunction (see Supplementary Table 3) and were excluded from Vestibular Agnosia (VA) group classification; VA-/VA+ analyses therefore include 155 participants.
Abbreviation: VOR, vestibulo-ocular reflex; MoCA, Montreal Cognitive Assessment; TMT-B, Trail Making Test – Part B; HADS-A and -D, Hospital Anxiety and Depression Scale – Anxiety and – depression; FES-I, Falls Efficacy Scale–International; TUG, Timed-Up-Go test; SD, standard deviation; IQR, interquartile range.

### Aim 1: Prevalence of Vestibular Agnosia (VA)

Among 166 older adults, 11 were classified as having peripheral vestibular hypofunction and were excluded from VA grouping (**Supplementary Table 3**). The remaining 155 with normal VOR thresholds was used to classify VA. An initial clustering of age and average perceptual thresholds in the full sample (young and older adults) yielded two clear clusters (silhouette coefficient: 0.69) (**Figure 3**): (1) a low-threshold group consisting of young adults (all 20 younger) and older adults (126/155 participants; 81.3%) and (2) a high-threshold older adult group (termed “vestibular agnosia”; 29/155; 18.7%).

**Figure 3.**
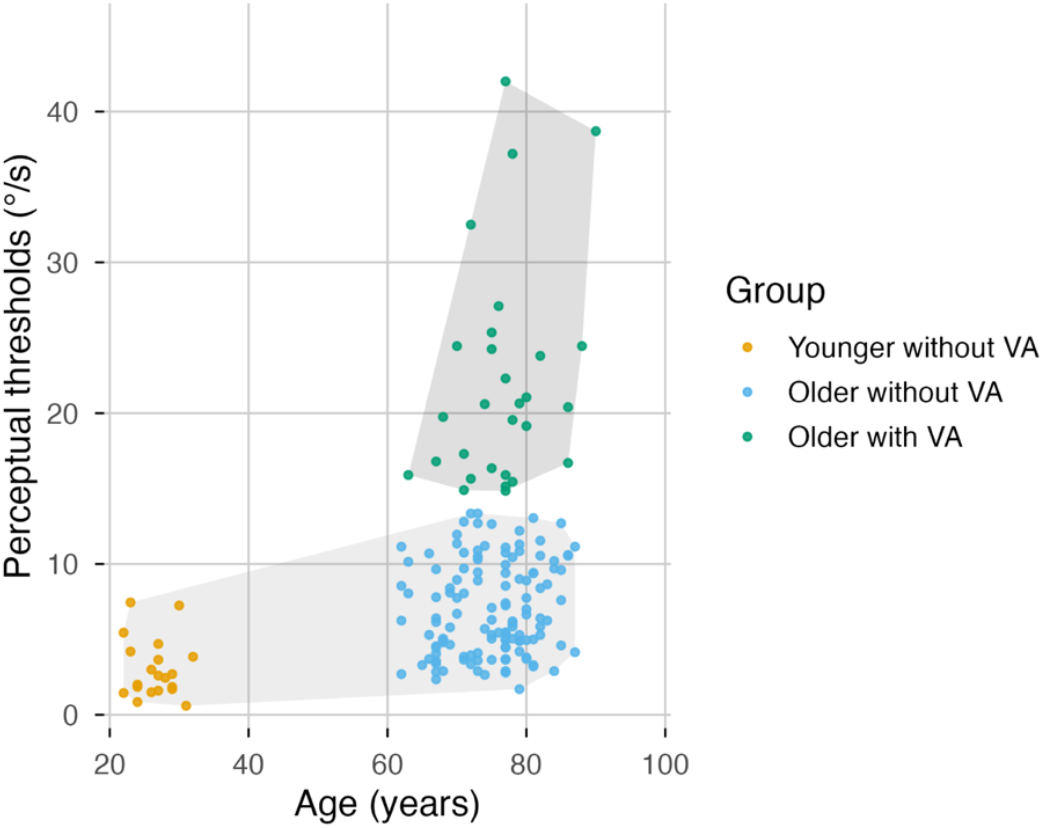
K-means clustering of age and average vestibular perceptual thresholds in younger (n=20) and older participants (n=155), classifying vestibular agnosia.

Participants with VA had – by definition – significantly higher perceptual thresholds (median: 20.4°/s vs. 6.3°/s). Although we excluded participants with peripheral vestibular dysfunction (VOR thresholds >9.65°/s), participants classified as VA+ nonetheless had slightly elevated VOR thresholds—albeit within “normal” bounds (median: 5.4°/s vs. 3.3°/s, *p*=0.006, **Table 1**). In unadjusted exploratory comparisons of baseline characteristics, participants with VA+ had reduced ankle vibration thresholds (*p*=0.02), longer sit-to-stand duration (*p*=0.03), lower cognitive function (MoCA) (*p*=0.01) and higher anxiety levels (*p*=0.04). Importantly, hand reaction time did not significantly differ between groups (*p*=0.89), indicating that poorer vestibular perception was not driven by reduced vigilance or slowed responding. Similarly, there were no significant differences in age, sex, executive function (TMT-B), or other demographic/health variables (**Table 1**).

### Aim 2: Vestibular Perceptual Thresholds and Balance

Among 159 participants with complete balance data, multivariable linear regression showed that higher (i.e. worse) perceptual thresholds independently related to both larger sway velocity (unadjusted B, 95% CI = 0.003, 0.001 to 0.004, *p*=0.003; adjusted B, 95% CI = 0.002, 0 to 0.004, *p*=0.03; **Figure 4A**) and area when standing with eyes-open on foam (unadjusted B, 95% CI = 0.002, 0.001 to 0.004, *p*<0.001; adjusted B, 95% CI = 0.002, 0 to 0.003, *p*=0.04; **Supplementary Table 1**). Under eyes-closed foam, perceptual thresholds were associated with greater sway in unadjusted analyses (Velocity: B, 95% CI = 0.007, 0.001 to 0.013, *p*=0.02; Area: B, 95% CI = 0.009, -0.022 to 0.040, *p*=0.58), but not after adjustment (Velocity: adjusted B, 95% CI = 0.003, −0.004 to 0.009, *p*=0.43; Area: adjusted B, 95% CI = -0.019, −0.052 to 0.015, *p*=0.28) (**Figure 4B** and **Supplementary Table 1**).

**Figure 4.**
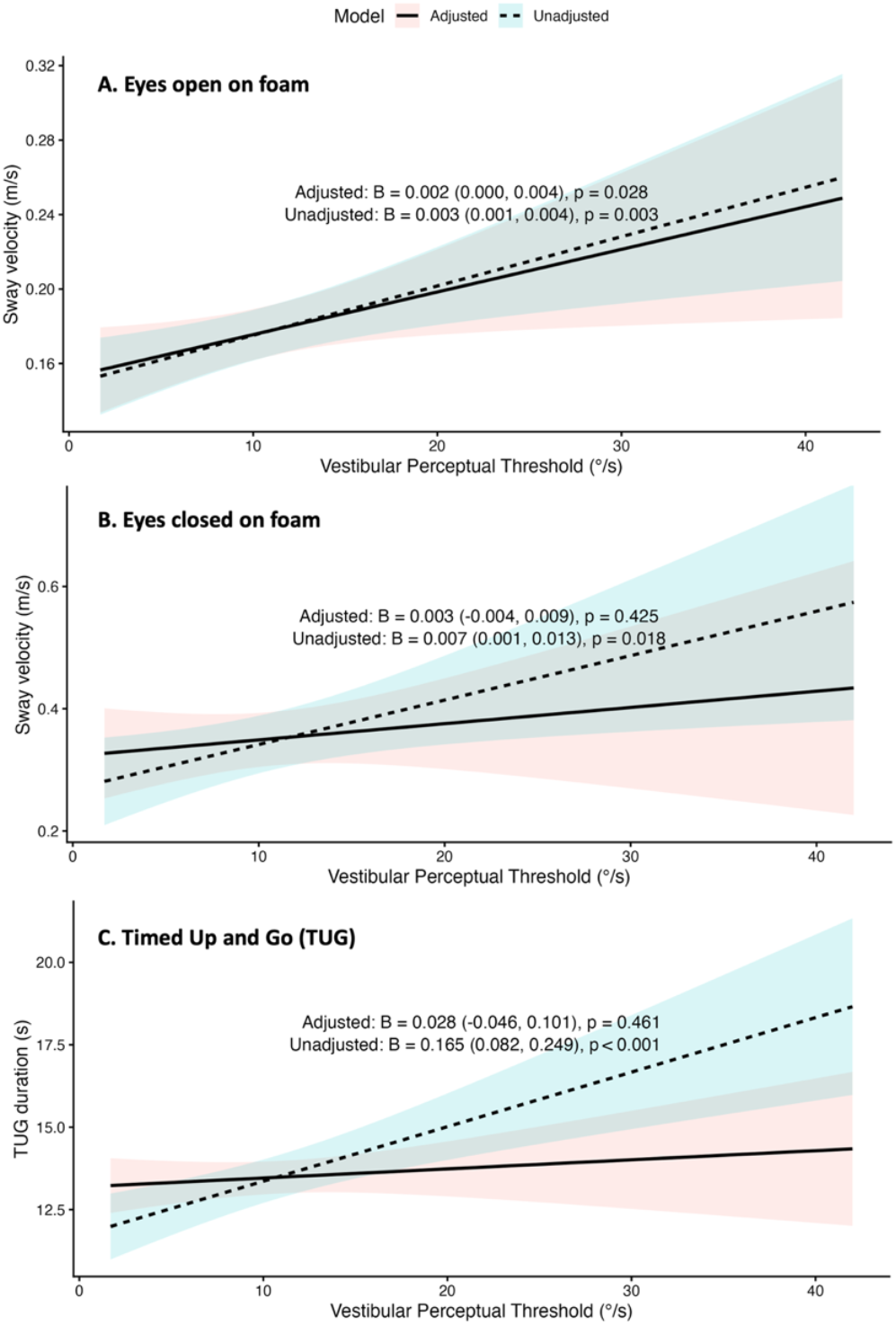
Linear regression of vestibular perceptual thresholds with sway velocity and Timed Up and Go performance. Predicted values from linear regression of vestibular perceptual threshold against **(A)** Sway velocity during eyes-open on foam; **(B)** Sway velocity during eyes-closed on foam; and **(C)** Timed Up and Go (TUG) duration. Solid lines show models adjusted for age, sex, MoCA, vestibulo-ocular reflex (VOR) thresholds, ankle vibration thresholds, and 5-times sit-to-stand; dashed lines show unadjusted models. Shaded areas represent 95% confidence intervals.

Perceptual thresholds were positively associated with longer TUG duration in unadjusted analysis (B, 95% CI = 0.165, 0.082 to 0.249, *p*<0.001), but not after adjustment (adjusted B, 95% CI = 0.028, −0.046 to 0.101, *p*=0.46) (**Figure 4C**). Perceptual thresholds were not associated with turn angle or velocity in either model (*p*>0.15; **Supplementary Table 1**).

### Aim 3: Vestibular Perceptual Thresholds and Prospective Falls

In the 161 participants (97%) who completed the full 6-month follow-up, 48 (29.8%) reported falling at least once (≥ 1 falls). Among those who fell, 27.1% experienced recurrent falls (≥ 2 falls). Faller status did not differ on either perceptual (Median: non-faller 8.55°/s, faller 9.43°/s; *p*=0.66) nor VOR thresholds (Median: non-faller 4.14°/s, faller 4.50°/s; *p*=0.31). We further examined the distribution of fall prevalence between the VA+ and VA– groups (note: 11 participants with peripheral vestibular hypofunction were excluded). Among the 45 participants who reported a fall, 11 (24.4%) had vestibular agnosia, whereas 17 (16.2%) of the 105 non-fallers had vestibular agnosia (*p*=0.26; **Supplementary Figure 2**).

Higher perceptual thresholds were not significantly associated with prospective falls (**Supplementary Table 2**). Perceptual thresholds were not associated with falls in logistics regression (OR, 95% CI = 1.01, 0.97 to 1.06, *p*=0.58; adjusted OR, 95% CI = 0.99, 0.94 to 1.04, *p*=0.65), nor with fall counts in negative binomial models (IRR, 95% CI = 1.03, 0.99 to 1.06, *p*=0.11; adjusted IRR, 95% CI = 1.02, 0.97 to 1.08, *p*=0.35).

## DISCUSSION

Our novel findings reveal that ∼19% of community-dwelling older adults had vestibular agnosia. These individuals had poorer cognition, reduced lower limb somatosensation and higher anxiety. Higher (i.e. worse) vestibular perceptual thresholds were independently associated with greater imbalance when standing on foam with eyes-open. Whilst similar associations were initially observed for eyes-closed on foam and gait performance, they were not significant after adjustment for important confounds (e.g., VOR thresholds). Contrary to our predictions, vestibular perceptual thresholds did not predict falls over 6-month.

### Prevalence of Vestibular Agnosia

Around one-fifth of our cohort of community-dwelling older adults exhibited vestibular agnosia. This is consistent with a recent study that found that ∼25% of older adults with BPPV reported absent dizziness [20]. Vestibular agnosia can increase missed inner-ear diagnoses (such as BPPV) by ∼7-10 times [17,44]. Diminished symptom awareness may limit help-seeking and adaptive safety behaviours (e.g., rising slowly or limiting rapid head movements), thereby potentially increasing fall risk. Future research should explore the specific consequences of vestibular agnosia in older adults with peripheral vestibular disorders, including BPPV whereby the link between vestibular agnosia and fall risk may be more apparent.

Individuals with vestibular agnosia exhibited lower global cognitive performance (MoCA) in exploratory between-group comparisons. Prior research has reported similarly modest links between perceptual thresholds and specific cognitive domains (attention, fluency, and language) in patients with traumatic brain injury [17]. Interestingly, vestibular agnosia did not relate to either executive function or vigilance in the present study, suggesting a more generalised and broad cognitive decline rather than a domain-specific deficit. This is consistent with neuroimaging evidence linking vestibular agnosia and related imbalance in patients with traumatic brain injury to bihemispheric fronto-posterior white matter network disruption [16].

### Vestibular perceptual thresholds and balance

Elevated (i.e. worse) perceptual thresholds were associated with increased sway velocity when standing on foam with eyes-open. These associations persisted even after adjustment for important covariates (including VOR thresholds, age, sex, MoCA, ankle vibration thresholds, and lower limb strength). This aligns with the prior studies linking poorer vestibular perception to increased postural sway [9–13]. However, unlike this prior work, our study isolates perceptual from peripheral deficits, highlighting the independent contributions of vestibular perception to balance in older adults.

Surprisingly, we found that perceptual thresholds were not associated with balance during the most challenging condition in which vestibular contributions to balance are maximized (eyes-closed on foam), following adjustments for covariates. Perceptual thresholds were independently associated with sway only under the eyes-open foam condition. Previous work in middle-aged traumatic brain injury patients had reported increased postural sway in individuals with vestibular agnosia standing on foam with both eyes open and closed [17]. Discrepancies may reflect cohort and task-related factors: eyes-closed foam standing may be disproportionately difficult for many older participants, limiting power to detect additional contributions of perceptual thresholds after covariate adjustment. Also, evidence linking perceptual thresholds to postural sway in older adults is mixed [9,10]. These discrepancies may reflect differences in the vestibular stimulus planes, sway metrics, and covariates adjustment (most of studies also did not control for peripheral vestibular function).

We focused on yaw-plane perceptual thresholds, given their relevance to turning control and turning-related fall risk, and prior evidence linking yaw-axis perception to clinically meaningful outcomes in other populations with vestibular agnosia (e.g., traumatic brain injury [17]). Mechanistically, on compliant surface the nervous system reweights sensory input from unreliable proprioception toward visual and vestibular cues [45]. The fact that yaw-plane perception related to sway observed only when vision was available alongside vestibular cues may suggest a specific role for such perception in visual-vestibular integration. By contrast, with vision removed, balance relies more on canal-otolith signals (measured by roll tilt thresholds) to maintain anteroposterior and mediolateral stabilization [10]. Yaw canal cues may still contribute by detecting and correcting unintended axial rotation, but contribute less direct information for gravitational tilt and stabilization, potentially explaining less variance in postural sway under this condition after covariates adjustment.

Although yaw-plane perceptual thresholds are mechanistically relevant to turning control [24], we observed no associations between perception in this axis and turning metrics (turn angle or velocity). A possible explanation is that this task was performed with vision available, allowing visual feedback to compensate for impaired vestibular perception, thereby reducing reliance on vestibular self-motion cues and attenuating associations with turning metrics. Whilst we found that perceptual thresholds were strongly associated with TUG duration in an unadjusted model, but eliminated after full adjustment, likely because TUG is heavily influenced by lower-limb strength and somatosensation (which were both controlled for) [46], and can be compensated in a predictable, visually guided environment [47]. Future work should examine more challenging conditions, such as uneven surfaces walking or stepping over obstacles, or turning reduced visual input.

### Vestibular perceptual thresholds and fall risk over 6-month follow-up

Contrary to our hypothesis and the cross-sectional balance associations, perceptual thresholds were not associated with 6-month prospective fall risk. There are several reasons for this. Firstly, vestibular agnosia is not always linked to imbalance, and can be a purely perceptual deficit [17], with neuroimaging data revealing only partial overlaps between circuits for vestibular agnosia and postural control [18]. Therefore, “vestibular agnosia” may represent a mechanistically heterogeneous group, with some individuals experiencing predominantly perceptual deficits and others with combined perceptual and balance impairments, which could attenuate associations between perceptual thresholds, postural outcomes, and fall risk. Secondly, many factors affect falls [48], including activity level and its riskiness.

Our findings differ from La Scaleia et al. [11], who reported that elevated pitch/roll thresholds predicted 1-year falls in older adults with type 2 diabetes. This discrepancy may reflect cohort differences. Compared with our relatively healthy volunteers, older adults with type 2 diabetes typically have greater multisensory and metabolic impairments [49], and higher burden of brain disease (including small vessel disease and dementia) [50], potentially amplifying the functional impact of vestibular perceptual deficits on falls. Accordingly, associations between vestibular perception and prospective falls may be more detectable in older adults with greater sensory/balance vulnerability (e.g., BPPV or impaired vision). Another important difference from La Scaleia et al. [11] is the lack of peripheral vestibular function assessment. This is a crucial, as diabetes is commonly associated with peripheral vestibular dysfunction [51]. It is therefore entirely plausible that both the poorer vestibular perception and increased falls are driven by peripheral vestibular dysfunction alone. Second, we assessed yaw-plane thresholds, while Scaleia et al. examined pitch/roll thresholds that are more directly linked to postural control and falls [7,52]. Finally, their findings were derived from only nine fall events (9.5%), likely inflating effect estimates, whereas our higher fall rate (28.2%; 48 events) provide more stable estimates despite null associations. Nonetheless, our study was likely underpowered for small-to-moderate effects.

### Implications for practice

Vestibular perception testing may be a promising tool to characterise vestibular-related balance impairment [21]. Future work could test whether it adds value beyond conventional assessments for risk stratification or mechanistic phenotyping, particularly in older adults with lower cognition and greater postural sway. Given the practical challenges of routinely assessing vestibular agnosia with a rotating chair, preliminary evidence indicates a clinic-feasible alternative: jointly measuring perceptual and VOR responses during caloric testing [53,54]. However, future work is needed to determine the optimal measure and to confirm whether caloric-defined vestibular agnosia exhibits similar associations with behaviourally relevant outcomes as standardised chair testing.

## Conclusions

Vestibular agnosia was identified in ∼20% of healthy community-dwelling older adults, despite preserved peripheral function. Elevated vestibular perceptual thresholds were independently associated with greater sway velocity during eyes-open foam stance (and with slower TUG in unadjusted analyses). In contrast, perceptual thresholds did not predict 6-month prospective falls, highlighting the multifactorial nature of fall risk. Overall, these findings underscore the potential value of vestibular perceptual testing for assessing age-related balance impairments, although its utility in fall-risk prediction requires further investigation.

## Data Availability

All data produced in the present study are available upon reasonable request to the authors.

## Acknowledgements

We would like to express our heartfelt gratitude to all the participants for their time, dedication, and enthusiastic involvement in this study.

## Notes

**Funding:** This work was supported by Lee Family Scholarship awarded to Yuxiao Li and a Wellcome Trust Sir Henry Wellcome Postdoctoral Fellowship awarded to Toby J. Ellmers (Grant Number: 222747/Z/21/Z). Zaeem Hadi is funded by Imperial College NIHR BRC. Rebecca Smith, Senior Clinical and Practitioner Research Award, NIHR304608 is funded by the NIHR and Imperial NIHR College BRC. The views expressed are those of the authors and not necessarily those of the NIHR or the Department of Health and Social Care. Barry M. Seemungal is funded by Medical Research Council UK, Imperial NIHR College BRC, Moulton Foundation.

**Conflict of interest:** The authors have no competing interests to report.

### Competing Interest Statement

The authors have declared no competing interest.

### Funding Statement

This work was supported by Lee Family Scholarship awarded to Yuxiao Li and a Wellcome Trust Sir Henry Wellcome Postdoctoral Fellowship awarded to Toby J. Ellmers (Grant Number: 222747/Z/21/Z). Zaeem Hadi is funded by Imperial College NIHR BRC. Rebecca Smith, Senior Clinical and Practitioner Research Award, NIHR304608 is funded by the NIHR and Imperial NIHR College BRC. The views expressed are those of the authors and not necessarily those of the NIHR or the Department of Health and Social Care. Barry M. Seemungal is funded by Medical Research Council UK, Imperial NIHR College BRC, Moulton Foundation.

### Author Declarations

East of England Essex Research Ethics Committee of the Health Research Authority gave ethical approval for this work (IRAS ID: 324021).

