## Supplementary Methods, Tables and Figures for "Vestibular Perception, Balance Impairment, and Fall Risk in Community-Dwelling Older Adults"

- Vestibular perceptual and vestibulo-ocular reflex (VOR) thresholds
- Demographics and health-related variables
- Statistical Analysis

**Supplementary Table 1.** Association of vestibular perceptual thresholds with balance in Multivariable Linear Regression

**Supplementary Table 2.** Predictors and prospective falls: results from binary logistic and negative binomial regression models

**Supplementary Table 3.** Symptom profiles and fall outcome across vestibular subgroups

**Supplementary Figure 1.** K-means clustering of left and right vestibular perceptual thresholds in older adults

**Supplementary Figure 2.** Prevalence of falls over 6 months in participants with and without vestibular agnosia

### Supplementary Methods

#### Vestibular perceptual and vestibulo-ocular reflex (VOR) thresholds

**Figure 1** illustrates the apparatus used to objectively access vestibular perceptual thresholds and VOR thresholds.

Vestibular perceptual thresholds were assessed using a vibration-free motorized rotating chair (Contraves, USA), delivering passive yaw-plane rotations (Figure 1A) . Participants were seated upright and securely restrained using a safety harness, with an adjustable head support to minimize head movement. To eliminate non-vestibular motion cues, the chair was enclosed with blackout curtains, all lights were turned off, and white noise was provided via two speakers mounted on the chair. Lights were briefly turned on between rotations to avoid post-rotatory vestibular effects.

Participants were asked to press the button (left or right) as soon as they perceived a rotation in the respective direction, via a hand-held keypad (Figure 1A). Stimuli followed a constant acceleration profile, with acceleration determined by the modified binary search (MOBS) staircase algorithm. Test rotations were of constant angular acceleration up to a maximum of 5s (i.e. 0.2Hz), followed by an identical deceleration phase. Participant responses indicating the correct direction made within the 5s acceleration phase was logged as a 'correct' response. Wrong direction or late responses (i.e. after the 5s acceleration phase) was logged as a 'missed' or wrong response (Figure 1B). Independent staircase algorithms were used for leftward and rightward rotations in randomized order. Each staircase terminated when two criteria were met: (1) three response reversals, and (2) convergence of acceleration values within 5% of the stimulus range ($0.1°/s^2$ to $11.1°/s^2$). Perceptual thresholds were defined as the midpoint between the final supra-threshold and sub-threshold values for each direction. Vestibular perceptual thresholds were thus obtained for left and rightward, and an average of left and right was also obtained. More detailed information regarding the specific algorithm used can be found in our previous studies. As with prior work [17, 27], perceptual threshold values were converted into velocity (°/s).

VOR thresholds were obtained simultaneously during perceptual thresholds measurements. However, when perceptual thresholds were much higher than VOR thresholds, the algorithm rapidly moved away from the peri-VOR thresholds range. Therefore, we additionally assessed VOR thresholds using the same chair apparatus via a previously used protocol whereby participants underwent yaw-plane rotations with linearly increasing acceleration ($0.3°/s^2$ every 3 seconds). Each trial ended upon a correct directional response (button press) or after 33 seconds if no response was logged (maximum velocity: 59.4 °/s). Each direction was tested twice. Eye movements were acquired using DC-coupled horizontal electrooculography (EOG; electrodes placed on the lateral canthi of each eye) acquired at a sampling rate of 250 Hz. Participants were instructed to look straight ahead during the testing. In line with previous work, the VOR threshold (°/s) was defined as the chair angular velocity at the onset of first consistent deviation in EOG signal from baseline (i.e., slow-phase eye movement; Figure 1B), followed by two clearly identifiable slow-phase/fast-phase nystagmus complexes. VOR thresholds were independently analysed by two authors (Y.L and T.J.E.). Any disagreements with discussed with a third author (R.M.S.).

To control for potential confounding effects of slowed reaction speed and vigilance on vestibular perceptual thresholds, participants also completed a simple computerised hand reaction time task. They were instructed to press a left or right button in response to congruent directional arrows presented on a laptop screen. Mean reaction time across correct responses was calculated.

**Demographics and health-related variables**

Baseline demographic and medical history included age, sex, ethnicity (white *vs.* non-white), educational level, diagnosis of hypertension and diabetes, number of medications (≥ 4 or not), smoking history (smoke/quitted *vs.* never smoked), alcohol use, previous falls and dizziness history in last 12-month. Validated questionnaires assessed psychological function (Short Falls Efficacy Scale–International (FES-I); Hospital Anxiety and Depression Scale (HADS)), cognitive and executive function (Montreal Cognitive Assessment (MoCA); Trail Making Test Part B (TMT-B), alongside dizziness-related symptom burden and visually induced dizziness (Dizziness Handicap Inventory (DHI); Visual Vertigo Analogue Scale (VVAS)).

Anthropometric data included height, weight, body mass index (BMI). Lower limb strength and physical function was measured using an instrumented 5-times sit-to-stand test (calculated using the Mobility Lab algorithm described above). Ankle vibration thresholds (as a proxy for lower limb somatosensation) were measured using a calibrated tuning fork (64 HZ) at the medial malleolus. Whilst seated with eyes-closed, participants indicated when they perceived the vibration had stopped.

**Statistical Analysis**

Normality was examined with the Shapiro–Wilk test. Continuous variables are reported as means ± standard deviations (SD) or medians with interquartile ranges (IQR); and categorical variables as frequencies with percentages.

***Aim 1: Prevalence of vestibular agnosia in older adults.*** Participants with abnormal VOR thresholds were excluded from this first aim, given that vestibular agnosia refers to impaired vestibular perception despite intact peripheral function [18]. To ensure we included all older participants with intact VOR function, we implemented a two-step exclusion protocol based on VOR thresholds. First, we established a normative cutoff for vestibular perceptual thresholds using data from younger adults (n=20; mean ± SD: 3.04 ± 1.95°/s), defined as mean + 2 SDs: 6.94°/s. Next, we calculated the VOR threshold distribution for older adults (n=69) who met the normative vestibular perceptual thresholds criterion ($\leq 6.94°/s^2$). The mean (*M*= 4.17°/s) + 2 SDs (*SD*= 2.74°/s) of this group's VOR thresholds served as the threshold value for abnormal VOR function, that is 9.65°/s. Participants were classified as demonstrating peripheral vestibular hypofunction if both vestibular perceptual (>6.94°/$s^2$) and ocular thresholds (>9.65°/s) were abnormal, resulting in 11 participants (6.6% of the sample) being excluded from this first analysis. (Please see Supplementary Table 1 for further information on this 'Peripheral Vestibular Hypofunction' sample.)

K-means clustering was first applied to the full sample of younger and older adults, using age and the average of left- and rightward perceptual thresholds as input variables. The optimal number of clusters was selected using the silhouette index and inspection of the elbow plot. To examine whether similar data-driven subgroups could be identified within the older cohort, and to align with previous work that used direction-specific perceptual thresholds, we next repeated k-means clustering in older adults only, using leftward and rightward perceptual thresholds as the input variables to identify individuals with and without vestibular agnosia.

Due to the highly imbalanced group sizes, vestibular agnosia was not used as a categorical predictor in the subsequent regression analysis; instead, vestibular perceptual thresholds were analysed as a continuous predictor variable in analyses performed for Aims 2 and 3 (see below).

***Aim 2: Association between vestibular perceptual thresholds and balance.*** Multivariable linear regression models were constructed to examine the association between perceptual thresholds (independent variable) and each outcome separately, including postural sway parameters (sway velocity, sway area) and TUG performance metrics (turn velocity, total duration, turn angle). Unadjusted models were followed by fully adjusted models controlling for age, sex, MoCA score, VOR thresholds, ankle vibration thresholds, and 5 times sit-to-stand (lower limb strength/physical function). Covariates were prespecified based on known contributors to performance on these tasks, as well as based on variables associated with impaired vestibular perception in Aim 1. All regression assumptions were met. Residuals were normally distributed and homoscedastic, with no evidence of collinearity (all VIFs < 1.4). Influential outliers were examined using Cook's distance; no cases exerted undue influence on model estimates. Standardized β coefficients, unstandardized B with 95% confidence intervals (CI) and corresponding *p* values were reported.

***Aim 3: Vestibular perception and prospective falls.*** Here, two regression approaches were used. A binary logistic regression model was used to evaluate the association between vestibular perception and the odds of experiencing ≥1 fall during the 6-month follow-up (yes/no), whilst a negative binomial regression was used to explore the predictors of total number of falls during follow-up period (fall counts). Because all participants had identical follow-up durations (6 months), no exposure offset term was included in the count models. For both outcomes, two models were applied. These were: (1) Unadjusted model, which included perceptual thresholds as the sole predictor (other covariates were also reported); (2) Adjusted model, which controlled for VOR thresholds, age, sex, fall history, ankle vibration thresholds, and lower-limb muscle strength (Sit-to-Stand duration). Model estimates are presented as odds ratios (OR), Incidence Rate Ratios (IRR) and 95% CI.

**Supplementary Table 1. Association of vestibular perceptual thresholds with balance in Multivariable Linear Regression (N=159)**

| Outcome | Condition | Unadjusted model | | *p* | Adjusted model | | *p* |
|---|---|---|---|---|---|---|---|
| | | Std. β | B (95% CI) | | Std. β | B (95% CI) | |
| **Sway velocity** (m/s) | EO-Foam | 0.233 | 0.003 (0.001, 0.004) | **0.003** | 0.203 | 0.002 (0.000, 0.004) | **0.03** |
| **Sway area** ($m^2/s^4$) | EO-Foam | 0.283 | 0.002 (0.001, 0.004) | **<0.001** | 0.180 | 0.002 (0.000, 0.003) | **0.04** |
| **Sway velocity** (m/s) | EC-Foam | 0.187 | 0.007 (0.001, 0.013) | **0.02** | 0.068 | 0.003 (-0.004, 0.009) | 0.43 |
| **Sway area** ($m^2/s^4$) | EC-Foam | 0.044 | 0.009 (-0.022, 0.040) | 0.58 | -0.094 | -0.019 (-0.052, 0.015) | 0.28 |
| **TUG duration** (s) | - | 0.299 | 0.165 (0.082, 0.249) | **<0.001** | 0.050 | 0.028 (-0.046, 0.101) | 0.46 |
| **Turn angle** (°) | - | -0.103 | -0.149 (-0.389, 0.090) | 0.220 | 0.004 | 0.006 (-0.248, 0.259) | 0.97 |
| **Turn velocity** (°/s) | - | -0.120 | -0.648 (-1.541, 0.245) | 0.154 | 0.093 | 0.504 (-0.303, 1.311) | 0.22 |

Abbreviation: EO, eyes open; EC, eyes closed; TUG, Timed-Up-Go test; CI, confidence interval.
The fully adjusted model was adjusted for age, sex, MoCA score, vestibulo-ocular reflex (VOR) thresholds, ankle vibration thresholds, and 5 times sit-to-stand.

**Supplementary Table 2. Predictors and prospective falls: results from binary logistic and negative binomial regression models (N = 161)**

| **Variable** | **Unadjusted model** | | **Adjusted model*** | |
|---|---|---|---|---|
| **Logistic regression (outcome: ≥1 fall)** | | | | |
| | **OR (95% CI)** | ***p*** | **OR (95% CI)** | ***p*** |
| Perceptual threshold (°/ s) | 1.01 (0.97, 1.06) | 0.58 | 0.99 (0.94, 1.04) | 0.65 |
| VOR threshold (°/ s) | 1.0 (0.95, 1.10) | 0.56 | 1.02 (0.94, 1.11) | 0.60 |
| Age (years) | 1.03 (0.98, 1.09) | 0.23 | 1.03 (0.97, 1.11) | 0.32 |
| Sex (female vs. male) | 0.88 (0.37, 2.19) | 0.77 | 1.05 (0.41, 2.91) | 0.91 |
| Past 12-month fall history | 1.69 (0.85, 3.36) | 0.13 | 1.29 (0.62, 2.68) | 0.50 |
| Ankle vibration threshold | 0.93 (0.78, 1.10) | 0.39 | 0.92 (0.75, 1.13) | 0.40 |
| 5 Sit to Stand duration (s) | 1.07 (0.99, 1.15) | 0.08 | 1.07 (0.99, 1.16) | 0.09 |
| **Negative binomial regression (outcome: fall count)** | | | | |
| | **IRR (95% CI)** | ***p*** | **IRR (95% CI)** | ***p*** |
| Perceptual threshold (°/ s) | 1.03 (0.99, 1.06) | 0.11 | 1.02 (0.97, 1.08) | 0.35 |
| VOR threshold (°/ s) | 1.00 (0.94, 1.07) | 0.90 | 0.98 (0.92, 1.05) | 0.66 |
| Age (years) | 1.04 (0.99, 1.09) | 0.14 | 1.02 (0.97, 1.07) | 0.47 |
| Sex (female vs. male) | 0.74 (0.37, 1.53) | 0.41 | 0.73 (0.33, 1.58) | 0.43 |
| Past 12-month fall history | 1.88 (1.07, 3.37) | 0.03 | 1.56 (0.84, 2.89) | 0.15 |
| Ankle vibration threshold | 0.90 (0.78, 1.04) | 0.15 | 0.94 (0.81, 1.09) | 0.41 |
| 5 Sit to Stand duration (s) | 1.02 (0.96, 1.08) | 0.56 | 1.00 (0.95, 1.06) | 0.89 |

*Adjusted for vestibulo-ocular reflex (VOR) thresholds, age and sex, fall history, ankle vibration threshold, and sit to stand duration.

Abbreviation: OR, odds ratio; IRR, incidence rate ratio; CI, confidence interval.

**Supplementary Table 3. Symptom profiles and fall outcome across vestibular subgroups (N=166)**

| Variable | Normal (n = 126) | VA (n = 29) | PVH (n = 11) | *p* |
|---|---|---|---|---|
| DHI score, median (IQR) | 0 (0, 0) | 0 (0, 1) | 0 (0, 0) | 0.17 |
| VVAS score, median (IQR) | 0 (0, 0) | 0 (0, 1) | 0 (0, 0) | 0.07 |
| Perceptual threshold (°/ s), median (IQR) | 6.28 (4.45, 9.70) | 20.40 (16.35, 24.45) | 17.65 (10.90, 28.80) | <0.001 |
| VOR threshold (°/ s), median (IQR) | 3.76 (2.31, 5.46) | 5.40 (3.78, 6.96) | 13.20 (12.45, 24.15) | <0.001 |
| Dizziness in past 12-month (yes), n (%) | 36 (29%) | 14 (48%) | 2 (18%) | 0.09 |
| Past 12-month fall history (yes), n (%) | 49 (40%) | 13 (46%) | 7 (70%) | 0.15 |
| Short FES-I, median (IQR) | 9 (8, 11) | 10 (8, 12) | 8 (8, 11) | 0.21 |
| HADS-A score, median (IQR) | 3 (1, 5) | 5 (2, 8) | 3 (2, 7) | 0.11 |
| TUG duration (s), median (IQR) | 12.45 (10.38, 14.54) | 12.98 (11.57, 15.04) | 13.45 (9.64, 19.48) | 0.33 |
| Turn velocity (°/s), median (IQR) | 173.93 (144.09, 204.71) | 170.40 (142.17, 192.51) | 154.02 (122.22, 173.94) | 0.19 |
| Sway velocity EC-Foam (m/s), median (IQR) | 0.16 (0.11, 0.20) | 0.18 (0.13, 0.25) | 0.19 (0.15, 0.25) | 0.10 |
| Fall in six-month follow-up (yes), n (%) | 34 (27.9%) | 11 (39.3%) | 3 (27.3%) | 0.48 |

Abbreviation: VA, Vestibular Agnosia; PVH, Peripheral Vestibular Hypofunction; DHI, Dizziness Handicap Inventory; VVAS, Visual Vertigo Analog Scale; VOR, Vestibulo-ocular reflex; FES-I, Falls Efficacy Scale–International; HADS-A, Hospital Anxiety and Depression Scale – Anxiety subscale; TUG, Timed Up and Go; EC-Foam, eyes-closed standing onn foam surface; IQR, Interquartile Range.

p values: Kruskal–Wallis test for continuous variables; Fisher's exact test for categorical variables.

**Supplementary Figure 1. K-means clustering of left and right vestibular perceptual thresholds in older adults (N = 155)**

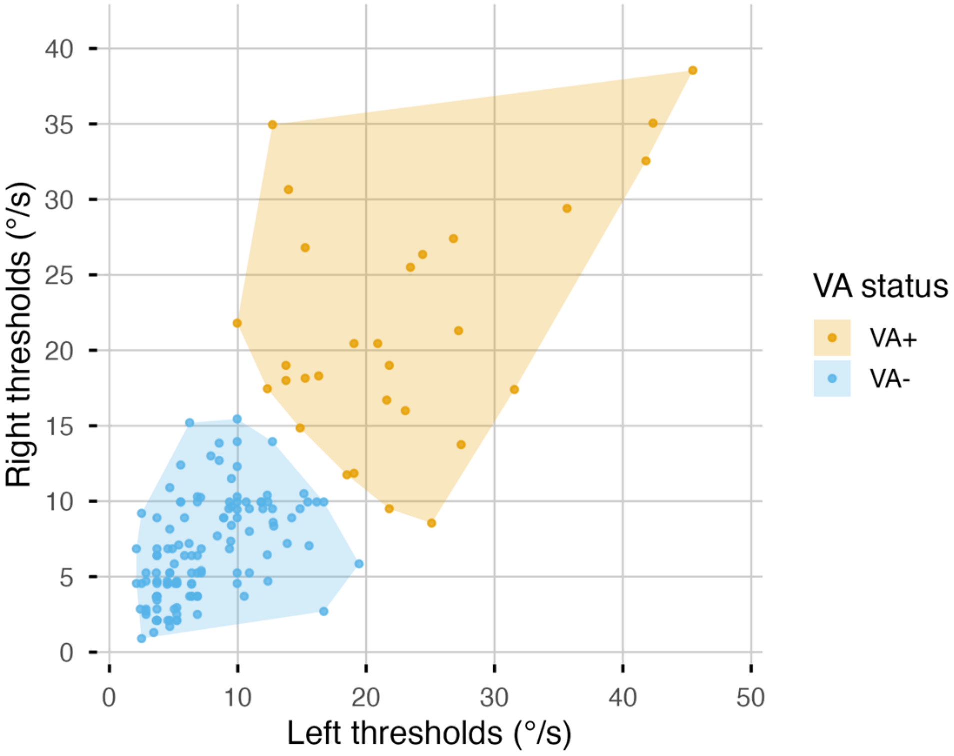


As a sensitivity analysis, K-means clustering of leftward and rightward perceptual thresholds in older adults identified two distinct clusters (VA+ vs. VA-). This matched the primary age – perceptual thresholds clustering approach (Figure 3).

**Supplementary Figure 2. Prevalence of falls over 6 months in participants with and without vestibular agnosia (N = 150)**

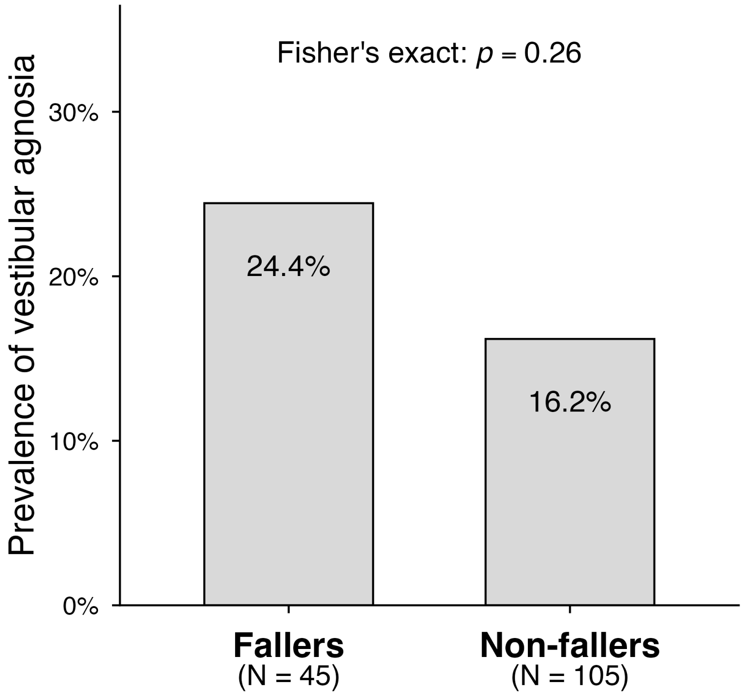
